# Resurgence of SARS-CoV-2 in England: detection by community antigen surveillance

**DOI:** 10.1101/2020.09.11.20192492

**Authors:** REACT Study Investigators:, Steven Riley, Kylie E. C. Ainslie, Oliver Eales, Caroline E. Walters, Haowei Wang, Christina Atchison, Claudio Fronterre, Peter J. Diggle, Deborah Ashby, Christl A. Donnelly, Graham Cooke, Wendy Barclay, Helen Ward, Ara Darzi, Paul Elliott

## Abstract

**Background:** Based on cases and deaths, transmission of SARS-CoV-2 in England peaked in late March and early April 2020 and then declined until the end of June. Since the start of July, cases have increased, while deaths have continued to decrease.

**Methods:** We report results from 594,000 swabs tested for SARS-CoV-2 virus obtained from a representative sample of people in England over four rounds collected regardless of symptoms, starting in May 2020 and finishing at the beginning of September 2020. Swabs for the most recent two rounds were taken between 24th July and 11th August and for round 4 between 22nd August and 7th September. We estimate weighted overall prevalence, doubling times between and within rounds and associated reproduction numbers. We obtained unweighted prevalence estimates by sub-groups: age, sex, region, ethnicity, key worker status, household size, for which we also estimated odds of infection. We identified clusters of swab-positive participants who were closer, on average, to other swab-positive participants than would be expected.

**Findings:** Over all four rounds of the study, we found that 72% (67%, 76%) of swab-positive individuals were asymptomatic at the time of swab and in the week prior. The epidemic declined between rounds 1 and 2, and rounds 2 and 3. However, the epidemic was increasing between rounds 3 and 4, with a doubling time of 17 (13, 23) days corresponding to an R value of 1.3 (1.2, 1.4). When analysing round 3 alone, we found that the epidemic had started to grow again with 93% probability. Using only the most recent round 4 data, we estimated a doubling time of 7.7 (5.5, 12.7) days, corresponding to an R value of 1.7 (1.4, 2.0). Cycle threshold values were lower (viral loads were higher) for rounds 1 and 4 than they were for rounds 2 and 3. In round 4, we observed the highest prevalence in participants aged 18 to 24 years at 0.25% (0.16%, 0.41%), increasing from 0.08% (0.04%, 0.18%) in round 3. We observed the lowest prevalence in those aged 65 and older at 0.04% (0.02%, 0.06%) which was stable compared with round 3. Participants of Asian ethnicity had elevated odds of infection. We identified clusters in and around London, transient clusters in the Midlands, and an expanding area of clustering in the North West and more recently in Yorkshire and the Humber.

**Interpretation:** Although low levels of transmission persisted in England through to mid-summer 2020, the prevalence of SARS-CoV-2 is now increasing. We found evidence of accelerating transmission at the end of August and beginning of September. Representative community antigen sampling can increase situational awareness and help improve public health decision making even at low prevalence.

## Introduction

SARS-CoV-2 infection continues to cause substantial COVID-19 morbidity and mortality globally [1]. Populations around the world are having to choose between higher levels of social contact [2] with higher levels of transmission versus lower levels of social contact and decreased levels of transmission [3]. This tradeoff also affects economic activity [4], non-COVID-19 related health and overall wellbeing. The ability of both individuals and governments to balance these competing demands requires accurate knowledge of trends in prevalence over time, person and place to enable informed choices to be made. COVID-19 surveillance systems in many countries are based on symptomatic cases, and are therefore subject to a number of biases, especially changes in care seeking behaviour that can obscure underlying trends [5].

We report here results of the REal-time Assessment of Community Transmission-1 (REACT-1) study, a large population-based programme designed to track prevalence of SARS-CoV-2 virus across England. We focus on rounds 3 and 4 conducted 24th July to 11th August and 22nd August to 7th September 2020 respectively, and compare these findings with those already reported from rounds 1 and 2 (1st May to 1st June and 19th June to 7th July 2020) [6,7].

## Methods

The protocol of the REACT suite of studies describes REACT-1 (antigen) and REACT-2 (antibody) [8]. Briefly, in REACT-1, we invited a random sample of the population aged five years and over, selected from the National Health Service (NHS) list of patients registered with a general practitioner, among the 315 lower-tier local authorities (LTLAs) in England. Among those registering to take part, in addition to age, sex, address and postcode available from the NHS register, we obtained, by online or telephone questionnaire, information on key worker status, ethnicity, smoking, household size, contact with known or suspected COVID-19 cases, hospital contacts, and symptoms. We provided written and video instructions to obtain a self-administered nose and throat swab (administered by a parent or guardian for children aged 5 to 12 years).

The four rounds of data collection obtained swab results on between 120,000 and 160,000 people at each round (Table 1). During the first round of data collection (1st May to 1st June 2020), swabs were initially collected in viral transport medium and sent to one of four Public Health England (PHE) laboratories for processing (n = 8,595 swabs with reported result). All subsequent collections were done using dry swabs which we requested that participants refrigerate before courier pick-up the same or next day. Swabs were then delivered to a commercial laboratory on a cold chain (4^0^ to 8^0^ C) to maintain sample integrity.

**Table 1.** Prevalence of swab-positivity across all four completed rounds of REACT-1.

| Round | First sample | Last sample | Positive | Total | Unweighted Prevalence (95% CI) | Weighted Prevalence (95% CI) |
| --- | --- | --- | --- | --- | --- | --- |
| 1 | 1/5/2020 | 1/6/2020 | 159 | 120,620 | 0.132% ( 0.113% , 0.154% ) | 0.156% ( 0.124% , 0.188% ) |
| 2 | 19/6/2020 | 7/7/2020 | 123 | 159,199 | 0.077% ( 0.065% , 0.092% ) | 0.088% ( 0.068% , 0.109% ) |
| 3 | 24/7/2020 | 11/8/2020 | 54 | 161,560 | 0.033% ( 0.026% , 0.044% ) | 0.040% ( 0.027% , 0.053% ) |
| 4 | 22/8/2020 | 07/09/2020 | 136 | 152,909 | 0.089% ( 0.075% , 0.105% ) | 0.126% ( 0.097% , 0.156% ) |

Samples were tested by reverse-transcription--polymerase-chain-reaction (RT-PCR) with two gene targets (E gene and N gene). We assessed cycle threshold (CT) values as a proxy for intensity of viral load across the four surveys. We defined swab to be positive if both gene targets were positive or if N gene was positive with CT value less than 37 [6].

### Analyses

To investigate trends in swab positivity over time, we used an exponential model of growth or decay, assuming that the number of positive samples each day arose from a binomial distribution. We used day of swabbing or, if unavailable, day of collection. Posterior credible intervals were obtained using a bivariate no-u-turn sampler [9] with uniform prior distributions. To estimate the reproduction number R, we assumed a generation time following a gamma distribution with a mean of 6.29 days and shape parameter 2.29 [10]. We analysed R using data from two sequential rounds and separately for each round. We fit an analogous model to the publicly available case data in which we assumed that the number of cases on a given day followed a negative binomial distribution. We estimated its dispersion parameter as an additional fitted parameter of the model.

We obtained crude prevalence estimates as the ratio of swab positive results to numbers tested for the total population and by sub-groups, e.g. age, sex, region, ethnicity, key worker status, household size. To correct for population differences due to sampling and differential response, we reweighted the oveall prevalence estimates to be representative of the English population by age, sex, region, ethnicity and deprivation. Because of small numbers of positive swabs, all other prevalence estimates were unweighted.

To investigate association of test result with covariates, we performed logistic regression adjusted for age and sex to obtain odds ratio estimates and 95% confidence intervals for each of the four rounds of data collection.

To investigate possible clustering of cases, we combined rounds 1 and 2, rounds 2 and 3, rounds 3 and 4, and calculated the distance between the home locations of every swab-positive participant, for which latitude and longitude were available, up to distances of 30 km. As a control, we randomly sampled the same number of swab-negative as swab-positive participants 5,000 times and calculated the equivalent distribution of distances.

We then identified individuals who appeared frequently within nearby pairs. Using samples of negative pairs, we obtained a null cumulative distribution of frequencies that participants appeared in pairs within 30 km of each other. We then constructed the corresponding single cumulative frequency curve for swab-positive participants. We defined a participant to be clustered if they appeared in nearby pairs more times than a minimum frequency, defined as that at which the swab-positive cumulative curve diverged from the central 90% region of the sampled swab-negative distribution. For our clustered swab-positive participants and 10,000 samples of the same number of swab-negative participants, we calculated the area of the convex hull which contained their home locations.

### Ethics

We obtained research ethics approval from the South Central-Berkshire B Research Ethics Committee (IRAS ID: 283787).

### Public involvement

A Public Advisory Panel is providing input into the design, conduct and dissemination of the REACT research programme.

## Results

In rounds 1 and 2 there was a downwards trend in prevalence [6,7] (Figure 1, Table 1). In round 3 we found 54 positive samples out of 161,560 swabs, giving an unweighted prevalence of 0.033% (95% CrI, 0.026%, 0.044%; Table 1) and a weighted prevalence of 0.040% (0.027%, 0.053%). Prevalence increased in round 4: we found 136 positive samples out of 152,909 swabs, giving an unweighted prevalence of 0.089% (0.075%, 0.105%) and a weighted prevalence of 0.126% (0.097%, 0.156%).

**Figure 1.**
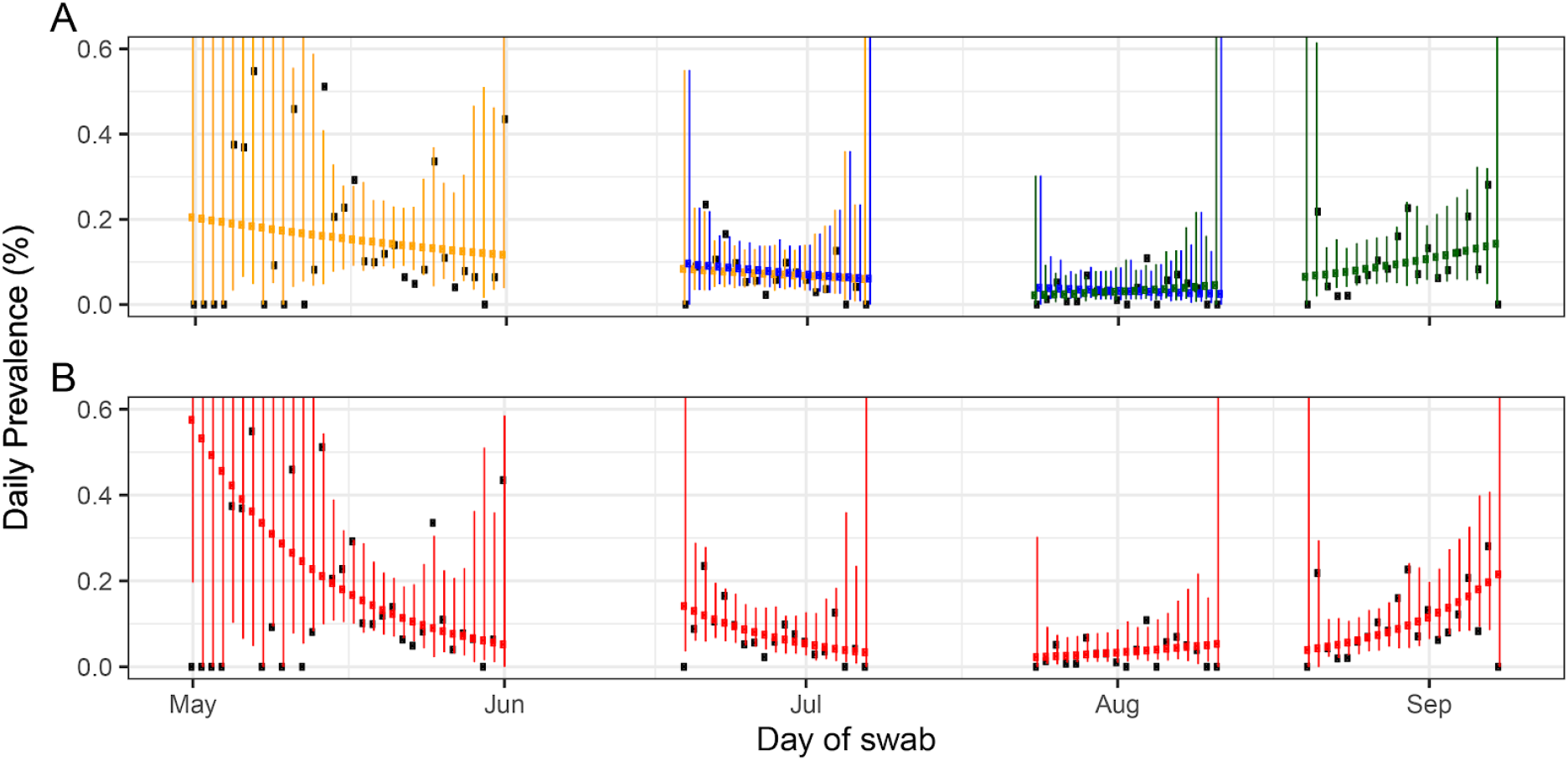
Constant growth rate models fit to REACT-1 data for sequential and individual rounds. **A** models fit to REACT-1 data for sequential rounds 1 and 2 (yellow), 2 and 3 (blue) and 3 and 4 (green). **B** models fit to individual rounds only (red). Vertical lines show 95% prediction intervals for models. Black points show observations. See Table 3 for R estimates.

Over all four rounds of the study, we found that 72% (67%, 76%) of swab-positive individuals were asymptomatic at the time of swab and in the week prior. Our data were suggestive of a higher rate of asymptomatic swab-positivity in children compared to adults (Table 2).

**Table 2.** Numbers and proportions of test positive children and adults by symptom status.

| Symptom status | Total |  | Children |  | Adults |  |
| --- | --- | --- | --- | --- | --- | --- |
|  | n | Proportion (95% CI) | n | Proportion (95% CI) | n | Proportion (95% CI) |
| No symptoms | 282 | 71.76% (67.11%, 75.98%) | 34 | 80.95% (66.70%, 90.01%) | 248 | 70.65% (65.69%, 75.17%) |
| Any symptoms | 111 | 28.24% (24.02%, 32.89%) | 8 | 19.05% (9.98%, 33.30%) | 103 | 29.34% (24.83%, 34.31%) |
| Classic COVID-19 symptoms* | 65 | 16.54% (13.19%, 20.53%) | 6 | 14.29% (6.72%, 27.84%) | 59 | 16.81% (13.26%, 21.06%) |

\* loss of sense of taste, loss of sense of smell, new persistent cough, fever

We estimated halving and doubling times and reproduction numbers across sequential rounds (Figure 1, Table 3). The epidemic declined between rounds 1 and 2, and between rounds 2 and 3. However, the epidemic increased between rounds 3 and 4, with a doubling time of 17 (13, 23) days corresponding to an R value of 1.3 (1.2, 1.4).

**Table 3.** Fitted growth rates, reproduction numbers and doubling times for SARS-CoV-2 swab positivity in England.

| Data | Round(s) | Number of participants / cases | Growth rate $r$ (1/days) | $P(r>0)$ | Reproduction number | Doubling (+) / halving (-) time (days) |
| --- | --- | --- | --- | --- | --- | --- |
| REACT All | 1 | 110,994 | -0.077 ( -0.107 , -0.046 ) | 0.000 | 0.6 ( 0.4 , 0.7 ) | -9.0 ( -6.5 , -14.9 ) |
|  | 2 | 157,428 | -0.089 ( -0.130 , -0.032 ) | 0.001 | 0.5 ( 0.4 , 0.8 ) | -7.8 ( -5.3 , -21.4 ) |
|  | 3 | 162,619 | 0.049 ( -0.012 , 0.109 ) | 0.937 | 1.3 ( 0.9 , 1.8 ) | 14.2 ( -58.6 , 6.4 ) |
|  | 4 | 152,554 | 0.090 ( 0.055 , 0.126 ) | 1.000 | 1.7 ( 1.4 , 2.0 ) | 7.7 ( 12.7 , 5.5 ) |
|  | 1 and 2 | 268,422 | -0.018 ( -0.025 , -0.012 ) | 0.000 | 0.9 ( 0.8 , 0.9 ) | -37.9 ( -28.0 , -57.5 ) |
|  | 2 and 3 | 320,047 | -0.025 ( -0.034 , -0.017 ) | 0.000 | 0.8 ( 0.8 , 0.9 ) | -27.3 ( -20.1 , -41.6 ) |
|  | 3 and 4 | 315,173 | 0.041 ( 0.030 , 0.052 ) | 1.000 | 1.3 ( 1.2 , 1.4 ) | 16.8 ( 23.0 , 13.2 ) |
| PHE Pillar 1 + 2 | 1 | 69,347 | -0.034 ( -0.042 , -0.027 ) | 0.000 | 0.8 ( 0.8 , 0.8 ) | -20.3 ( -16.7 , -26.0 ) |
|  | 2 | 11,550 | -0.018 ( -0.032 , -0.004 ) | 0.006 | 0.9 ( 0.8 , 1.0 ) | -38.6 ( -21.9 , -164.1 ) |
|  | 3 | 15,164 | 0.027 ( 0.005 , 0.049 ) | 0.990 | 1.2 ( 1.0 , 1.3 ) | 25.8 ( 137.9 , 14.2 ) |
|  | 4 | 21,277 | 0.058 ( 0.034 , 0.082 ) | 0.351 | 1.4 ( 1.2 , 1.6 ) | 11.9 ( 20.3 , 8.4 ) |
|  | 1 and 2 | 97,348 | -0.029 ( -0.032 , -0.027 ) | 0.000 | 0.8 ( 0.8 , 0.8 ) | -23.7 ( -22.0 , -25.7 ) |
|  | 2 and 3 | 36,420 | 0.007 ( 0.003 , 0.011 ) | 1.000 | 1.0 ( 1.0 , 1.1 ) | 94.0 ( 211.4 , 60.6 ) |
|  | 3 and 4 | 43,897 | 0.022 ( 0.015 , 0.029 ) | 0.997 | 1.1 ( 1.1 , 1.2 ) | 31.6 ( 45.3 , 24.2 ) |
| REACT, only double-gene positives count as positive (single count as negative) | 1 | 110,994 | -0.045 ( -0.093 , 0.005 ) | 0.047 | 0.7 ( 0.5 , 1.0 ) | -15.4 ( -7.4 , 141.9 ) |
|  | 2 | 157,428 | -0.112 ( -0.183 , -0.047 ) | 0.001 | 0.4 ( 0.2 , 0.7 ) | -6.2 ( -3.8 , -14.8 ) |
|  | 3 | 162,619 | 0.075 ( 0.000 , 0.149 ) | 0.969 | 1.5 ( 1.0 , 2.2 ) | 9.3 ( +Inf , 4.7 ) |
|  | 4 | 152,554 | 0.096 ( 0.049 , 0.142 ) | 0.982 | 1.7 ( 1.3 , 2.1 ) | 7.2 ( 14.2 , 4.9 ) |
|  | 1 and 2 | 268,422 | -0.012 ( -0.021 , -0.003 ) | 0.003 | 0.9 ( 0.9 , 1.0 ) | -55.5 ( -32.4 , -199.8 ) |
|  | 2 and 3 | 320,047 | -0.020 ( -0.031 , -0.008 ) | 0.000 | 0.9 ( 0.8 , 0.9 ) | -35.3 ( -22.1 , -84.6 ) |
|  | 3 and 4 | 315,173 | 0.041 ( 0.027 , 0.055 ) | 0.993 | 1.3 ( 1.2 , 1.4 ) | 17.1 ( 25.7 , 12.7 ) |
| REACT, Only asymptomatic positives count as positives | 1 | 110,968 | -0.095 ( -0.134 , -0.053 ) | 0.000 | 0.5 ( 0.3 , 0.7 ) | -7.3 ( -5.2 , -13.0 ) |
|  | 2 | 157,416 | -0.109 ( -0.169 , -0.053 ) | 0.000 | 0.4 ( 0.2 , 0.7 ) | -6.4 ( -4.1 , -13.2 ) |
|  | 3 | 162,606 | 0.069 ( -0.011 , 0.153 ) | 0.946 | 1.5 ( 0.9 , 2.2 ) | 10.1 ( -61.4 , 4.5 ) |
|  | 4 | 152,533 | 0.071 ( 0.023 , 0.119 ) | 0.995 | 1.5 ( 1.2 , 1.9 ) | 9.7 ( 30.2 , 5.8 ) |
|  | 1 and 2 | 268,384 | -0.013 ( -0.021 , -0.005 ) | 0.001 | 0.9 ( 0.9 , 1.0 ) | -54.0 ( -33.7 , -135.9 ) |
|  | 2 and 3 | 320,022 | -0.034 ( -0.047 , -0.023 ) | 0.000 | 0.8 ( 0.7 , 0.9 ) | -20.2 ( -14.9 , -30.2 ) |
|  | 3 and 4 | 315,139 | 0.040 ( 0.026 , 0.055 ) | 0.995 | 1.3 ( 1.2 , 1.4 ) | 17.4 ( 27.1 , 12.6 ) |

Within each round, swabs were collected over a sufficiently long period that we could also estimate rates of change in prevalence using data only from a single round. The epidemic was declining within rounds 1 and 2 (Figure 1, Table 3). However, we found evidence when analysing round 3 alone that the epidemic had started to grow again with a doubling time of 14 days and a 95% credible interval from halving every 59 days to doubling every 6.4 days, corresponding to an R value of 1.3 (0.9, 1.8) with a 93% probability that the R value was greater than 1. In round 4, we estimated that the doubling time had reduced to 7.7 (5.5, 13) days, corresponding to an R value of 1.7 (1.4, 2.0) with a probability greater than 99% that R was greater than 1 (Table 3).

In sensitivity analyses, our estimates of R and doubling times were similar when considering two relevant subsets of the swab-positivity variable (Table 3). For example, there were no changes to the estimate of 1.3 for R based on rounds 3 and 4 when we used data only from participants who did not report symptoms, and also when we considered positive samples to be only those where both gene targets were detected. Under these same scenarios, our estimates of R in round 4 only were 1.7 and 1.5 respectively, compared with 1.7 in the primary analysis.

At the regional scale, we had power to detect patterns of growth or decline when analysing rounds 3 and 4 together. Across these two rounds, there was positive growth in the epidemic in all regions (slightly lower confidence for East Midlands, Table 4, Figure 2). The highest growth across rounds 3 and 4 was in the North East with an R value of 1.7 (1.2, 2.5). During round 4, prevalence was highest in the North West (0.17%), Yorkshire and the Humber (0.17%), and North East (0.16%) (Table 5): participants in these three regions had 2.8- to 3-fold increased odds of infection relative to those in the South East (Table 6).

**Table 4.** Fitted growth rates, reproduction numbers and doubling times for SARS-CoV-2 swab positivity in regions of England.

| Region | Rounds | Number of participants / cases | Growth rate $r$ (1/days) | $P(r>0)$ | Reproduction number | Doubling (+) / halving (-) time (days) |
| --- | --- | --- | --- | --- | --- | --- |
| South East | 1 and 2 | 59,471 | -0.027 ( -0.041 , -0.014 ) | 0.00 | 0.8 ( 0.8 , 0.9 ) | -25.5 ( -16.7 , -50.9 ) |
|  | 2 and 3 | 70,437 | -0.033 ( -0.060 , -0.010 ) | 0.00 | 0.8 ( 0.7 , 0.9 ) | -20.9 ( -11.6 , -68.6 ) |
|  | 3 and 4 | 70,580 | 0.045 ( 0.017 , 0.077 ) | 1.00 | 1.3 ( 1.1 , 1.6 ) | 15.3 ( 40.8 , 9.0 ) |
| North East | 1 and 2 | 9,913 | -0.052 ( -0.119 , -0.004 ) | 0.02 | 0.7 ( 0.4 , 1.0 ) | -13.3 ( -5.8 , -179.4 ) |
|  | 2 and 3 | 11,586 | 0.0123 ( -0.08 , 0.117 ) | 0.61 | 1.1 ( 0.5 , 1.9 ) | 56.3 ( -8.22 , 5.936 ) |
|  | 3 and 4 | 11,450 | 0.0938 ( 0.033 , 0.177 ) | 1.00 | 1.7 ( 1.2 , 2.5 ) | 7.4 ( 21.13 , 3.927 ) |
| North West | 1 and 2 | 32,745 | -0.021 ( -0.04 , -0.004 ) | 0.01 | 0.9 ( 0.8 , 1.0 ) | -32.8 ( -17.9 , -158 ) |
|  | 2 and 3 | 37,946 | -0.017 ( -0.041 , 0.005 ) | 0.07 | 0.9 ( 0.8 , 1.0 ) | -40.4 ( -16.8 , 129.5 ) |
|  | 3 and 4 | 36,542 | 0.054 ( 0.028 , 0.080 ) | 1.00 | 1.4 ( 1.2 , 1.6 ) | 12.9 ( 24.5 , 8.6 ) |
| Yorkshire | 1 and 2 | 17,381 | -0.011 ( -0.041 , 0.019 ) | 0.23 | 0.9 ( 0.8 , 1.1 ) | -61.9 ( -16.8 , 35.8 ) |
|  | 2 and 3 | 21,089 | 0.001 ( -0.032 , 0.035 ) | 0.53 | 1.0 ( 0.8 , 1.2 ) | 547.9 ( -21.9 , 19.8 ) |
|  | 3 and 4 | 20,916 | 0.039 ( 0.009 , 0.073 ) | 1.00 | 1.3 ( 1.1 , 1.5 ) | 17.8 ( 76.8 , 9.5 ) |
| East Midlands | 1 and 2 | 34,757 | -0.021 ( -0.041 , -0.003 ) | 0.01 | 0.9 ( 0.8 , 1.0 ) | -32.4 ( -16.9 , -227.3 ) |
|  | 2 and 3 | 41,418 | -0.015 ( -0.039 , 0.008 ) | 0.11 | 0.9 ( 0.8 , 1.1 ) | -47.5 ( -17.7 , 82.5 ) |
|  | 3 and 4 | 40,137 | 0.017 ( -0.014 , 0.050 ) | 0.85 | 1.1 ( 0.9 , 1.3 ) | 40.6 ( -49.2 , 14.0 ) |
| West Midlands | 1 and 2 | 25,135 | -0.020 ( -0.039 , -0.001 ) | 0.02 | 0.9 ( 0.8 , 1.0 ) | -35.0 ( -17.7 , -679.6 ) |
|  | 2 and 3 | 29,965 | -0.032 ( -0.066 , -0.004 ) | 0.01 | 0.8 ( 0.6 , 1.0 ) | -21.5 ( -10.6 , -170.9 ) |
|  | 3 and 4 | 29,470 | 0.028 ( -0.008 , 0.067 ) | 0.94 | 1.2 ( 1.0 , 1.5 ) | 24.4 ( -87.6 , 10.4 ) |
| East of England | 1 and 2 | 39,008 | -0.013 ( -0.030 , 0.005 ) | 0.08 | 0.9 ( 0.8 , 1.0 ) | -53.6 ( -22.8 , 135.5 ) |
|  | 2 and 3 | 46,419 | -0.030 ( -0.057 , -0.005 ) | 0.01 | 0.8 ( 0.7 , 1.0 ) | -23.2 ( -12.1 , -138.0 ) |
|  | 3 and 4 | 46,259 | 0.047 ( 0.015 , 0.083 ) | 1.00 | 1.3 ( 1.1 , 1.6 ) | 14.9 ( 45.2 , 8.3 ) |
| London | 1 and 2 | 25,171 | -0.012 ( -0.028 , 0.004 ) | 0.06 | 0.9 ( 0.8 , 1.0 ) | -55.7 ( -24.4 , 186.6 ) |
|  | 2 and 3 | 30,550 | -0.034 ( -0.060 , -0.011 ) | 0.00 | 0.8 ( 0.7 , 0.9 ) | -20.5 ( -11.6 , -60.4 ) |
|  | 3 and 4 | 29,798 | 0.036 ( 0.006 , 0.070 ) | 0.99 | 1.2 ( 1.0 , 1.5 ) | 19.2 ( 122.7 , 9.9 ) |
| South West | 1 and 2 | 24,841 | -0.001 ( -0.025 , 0.023 ) | 0.46 | 1.0 ( 0.8 , 1.2 ) | -614.2 ( -27.3 , 29.6 ) |
|  | 2 and 3 | 30,637 | -0.058 ( -0.107 , -0.021 ) | 0.00 | 0.7 ( 0.4 , 0.9 ) | -12.0 ( -6.5 , -33.3 ) |
|  | 3 and 4 | 30,021 | 0.053 ( 0.010 , 0.109 ) | 0.99 | 1.4 ( 1.1 , 1.8 ) | 13.0 ( 69.5 , 6.4 ) |

**Figure 2.**
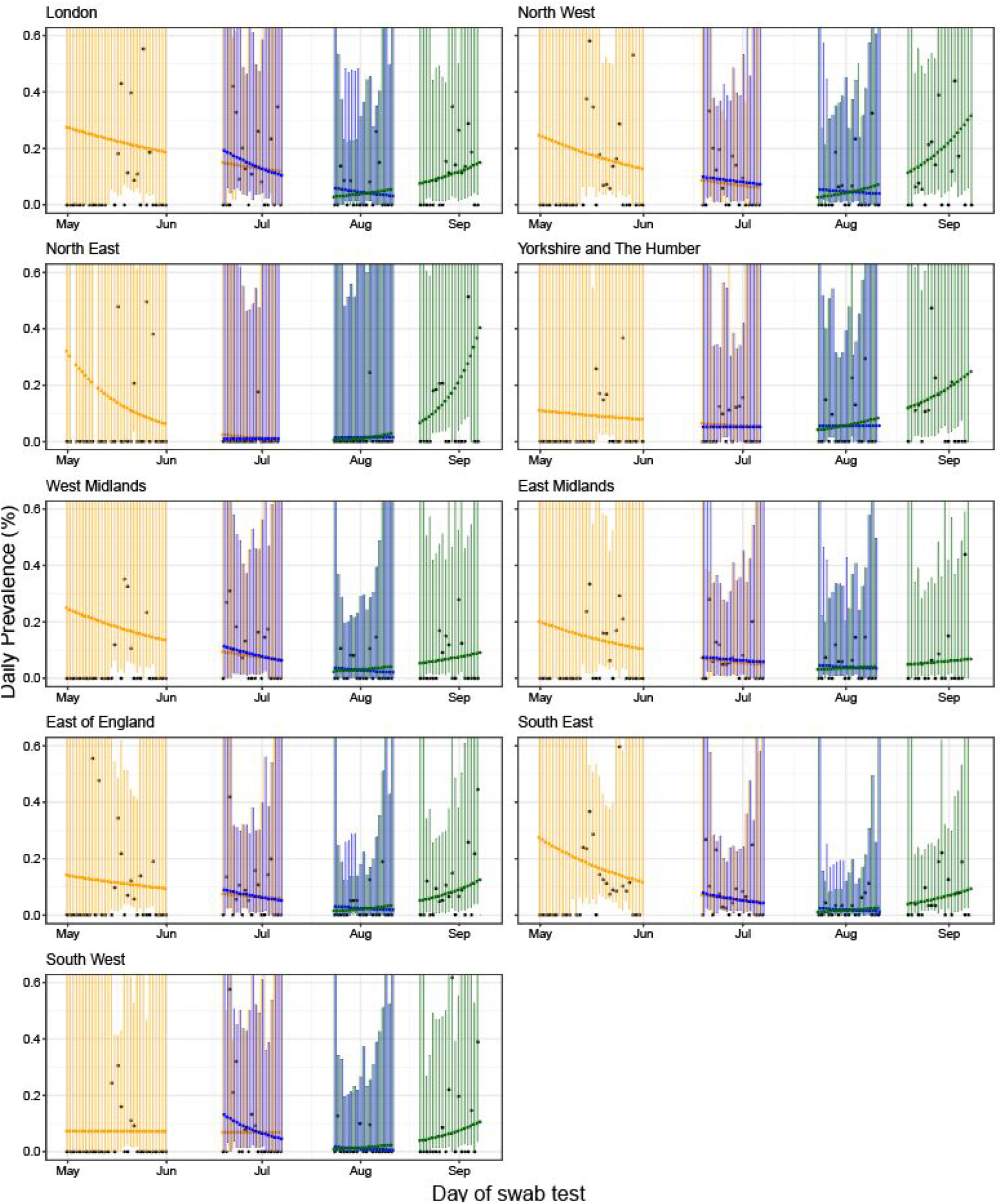
Constant growth rate models fit to sequential rounds of REACT-1 data by region. Models fit to rounds 1 and 2 (yellow), 2 and 3 (blue) and 3 and 4 (green). Vertical lines show 95% prediction intervals for models. Black points show observations. See Table 3 for associated R values.

In round 4, we observed highest prevalence in participants aged 18 to 24 years at 0.25% (0.16%, 0.41%) increasing from 0.08% (0.04%, 0.18%) in round 3 (Table 5). We observed the lowest prevalence in those aged 65 and older at 0.04% (0.02%, 0.06%) which was stable compared with round 3. The observed increase in prevalence in round 4 was not explained by health care and care home workers whose odds of infection were similar to those of other workers and had dropped substantially since round 1 (Table 6). Also, in round 4 participants of Asian ethnicity had odds of infection of 2.2 (1.2, 4.0) relative to whites (Table 6).

**Table 5.**
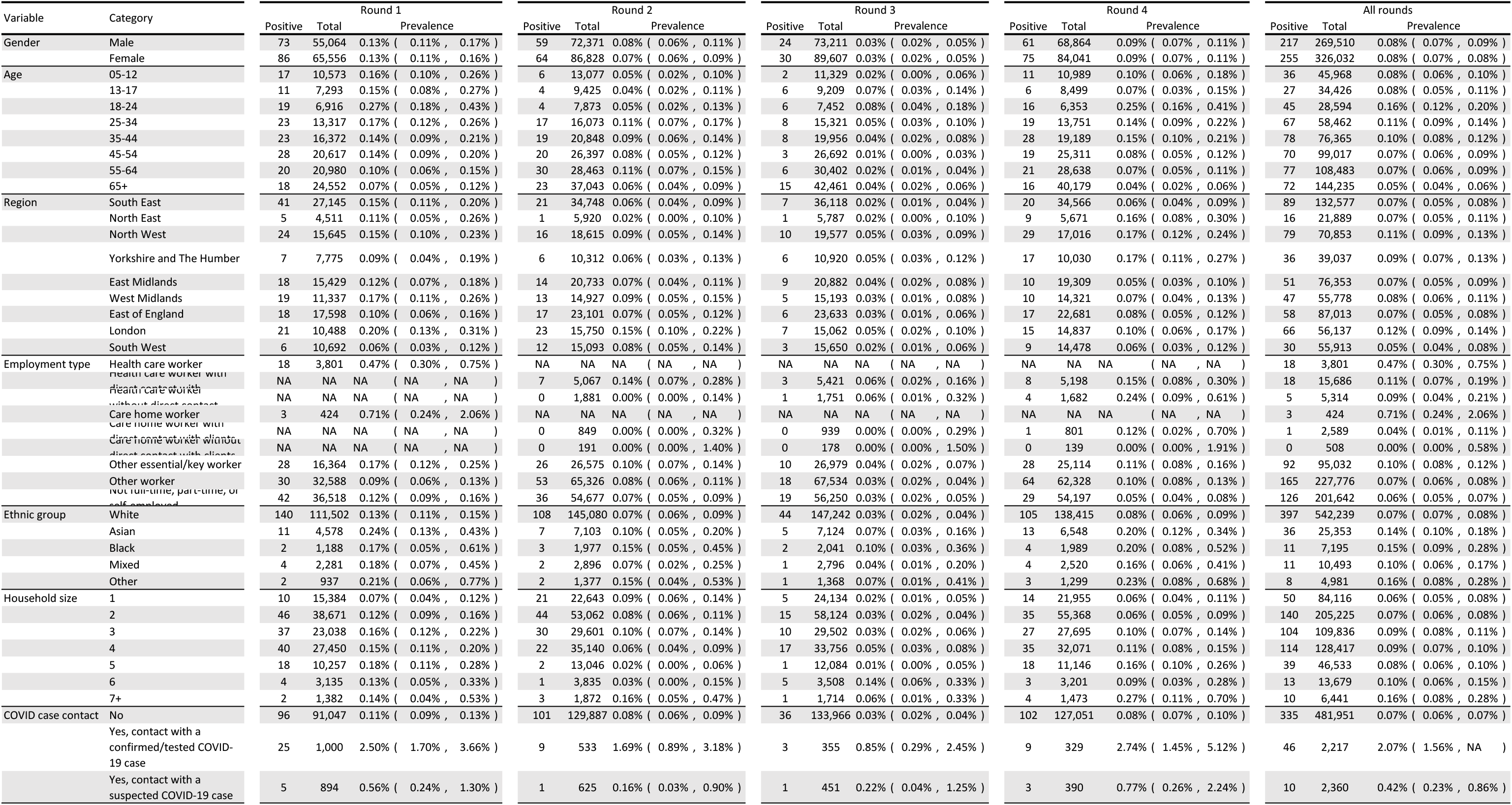
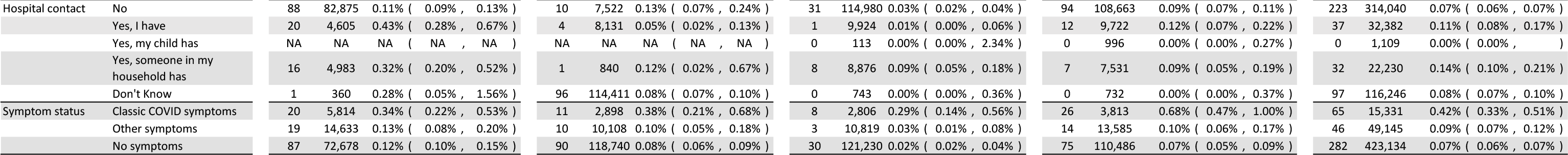
Prevalence of infection by round, variable and category. [Inserted on next 2 pages in pdf].

**Table 6.** Logistic regression models adjusted for age and gender for REACT-1 rounds 1, 2, 3 and 4.

| Variable | Category | Round 1 | Round 2 | Round 3 | Round 4 |
| --- | --- | --- | --- | --- | --- |
| Gender | Male | ref | ref | ref | ref |
|  | Female | 0.98 [0.71,1.33] | 0.90 [0.63,1.29] | 1.02 [0.60,1.75] | 0.98 [0.70,1.38] |
| Age group (yr) | 5 - 35 | 1.66 [1.15,2.38] | 0.79 [0.50,1.24] | 2.60 [1.24,5.46] | 1.29 [0.88,1.89] |
|  | 36 - 56 | ref | ref | ref | ref |
|  | 57 + | 0.73 [0.47,1.13] | 0.96 [0.64,1.45] | 1.61 [0.76,3.42] | 0.48 [0.30,0.75] |
| Region | South East | ref | ref | ref | ref |
|  | North East | 0.74 [0.29,1.87] | 0.28 [0.04,2.07] | 0.90 [0.11,7.28] | 2.78 [1.26,6.10] |
|  | North West | 1.01 [0.61,1.68] | 1.42 [0.74,2.73] | 2.62 [1.00,6.88] | 2.99 [1.69,5.29] |
|  | Yorkshire and The Humber | 0.60 [0.27,1.34] | 0.96 [0.39,2.38] | 2.83 [0.95,8.43] | 3.01 [1.57,5.74] |
|  | East Midlands | 0.77 [0.44,1.34] | 1.12 [0.57,2.20] | 2.23 [0.83,5.98] | 0.91 [0.42,1.94] |
|  | West Midlands | 1.10 [0.64,1.90] | 1.44 [0.72,2.88] | 1.69 [0.54,5.34] | 1.21 [0.57,2.59] |
|  | East of England | 0.67 [0.39,1.17] | 1.22 [0.64,2.31] | 1.31 [0.44,3.88] | 1.30 [0.68,2.48] |
|  | London | 1.24 [0.73,2.11] | 2.47 [1.37,4.48] | 2.30 [0.80,6.57] | 1.58 [0.81,3.10] |
|  | South West | 0.38 [0.16,0.89] | 1.31 [0.64,2.67] | 0.99 [0.26,3.83] | 1.12 [0.51,2.46] |
| Employment type | HCW / CHW | 5.49 [3.11,9.69] | 1.13 [0.51,2.50] | 1.85 [0.62,5.51] | 1.53 [0.84,2.79] |
|  | Other key worker | 1.83 [1.09,3.06] | 1.23 [0.76,1.97] | 1.44 [0.66,3.14] | 1.01 [0.65,1.58] |
|  | Other worker | ref | ref | ref | ref |
|  | Not FT, PT, SE | 1.40 [0.86,2.30] | 0.79 [0.50,1.25] | 1.16 [0.59,2.28] | 0.64 [0.40,1.01] |
| Ethnicity | White | ref | ref | ref | ref |
|  | Asian | 1.71 [0.92,3.17] | 1.37 [0.64,2.97] | 2.26 [0.88,5.76] | 2.21 [1.24,3.96] |
|  | Black | 1.26 [0.31,5.09] | 2.09 [0.66,6.59] | 3.20 [0.77,13.29] | 2.30 [0.85,6.27] |
|  | Mixed | 1.10 [0.40,2.99] | 1.02 [0.25,4.19] | 1.01 [0.14,7.46] | 1.64 [0.60,4.49] |
|  | Other | 1.63 [0.40,6.58] | 1.99 [0.49,8.08] | 2.44 [0.34,17.80] | 2.72 [0.86,8.61] |
| Household size | 1 - 2 | ref | ref | ref | ref |
|  | 3 - 5 | 1.14 [0.78,1.66] | 0.80 [0.53,1.22] | 1.71 [0.86,3.42] | 1.22 [0.81,1.84] |
|  | 6 + | 0.88 [0.40,1.90] | 0.83 [0.29,2.36] | 4.85 [1.76,13.42] | 1.53 [0.67,3.50] |

We compared epidemic trends estimated from REACT-1 data with those based on UK surveillance data for key workers (Pillar 1) and symptomatic individuals and those tested as part of a local response (Pillar 2) [11] (Table 3, Figure 3). Based on Pillar 1 and 2 data, using an exponential growth model, we found that numbers of cases were growing from 19th June to 11th August (start round 2 to end round 3), whereas our analysis of REACT-1 data for the same period found the epidemic to be declining. Results based on data since 24th July (start round 3), for both REACT-1 and Pillar 1 and 2 data showed a growing epidemic. However, the estimated rate of growth was higher in the REACT-1 data (Table 3).

**Figure 3.**
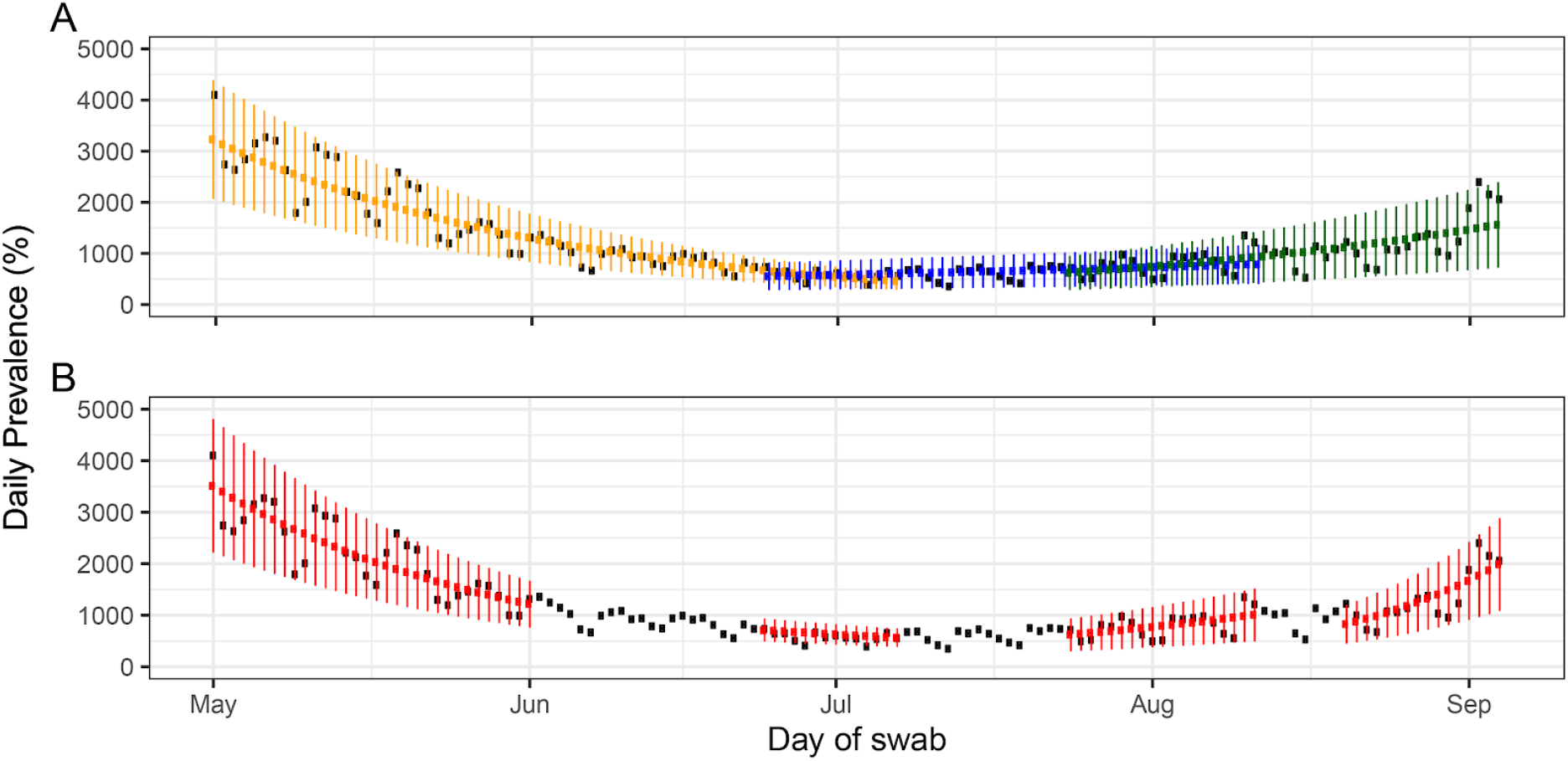
Constant growth rate models fit to UK government Pillar 1 and 2 data for sequential and individual rounds. **A** models fit to symptomatic and key worker test data for the periods of: rounds 1 and 2 (yellow), 2 and 3 (blue) and 3 and 4 (green). **B** models fit to the same period as each individual round (red). Note that only UK government data up to the 4^th^ September was included whereas the REACT data covered up until the 7^th^ September. This was done to avoid the problem of the most recent days in the UK government data having underreported values. Vertical lines show 95% prediction intervals for models. Black points show observations. See Table 3 for R estimates.

Cycle threshold values reflect the amount of virus in a positive sample, with high CT values corresponding to low levels of virus. There was a change in the distribution of CT values between rounds 1 and 2, not between rounds 2 and 3 but then again between rounds 3 and 4 (Figure 4, Table 7). CT values were lower (viral loads were higher) for rounds 1 and 4 than for rounds 2 and 3.

**Figure 4.**
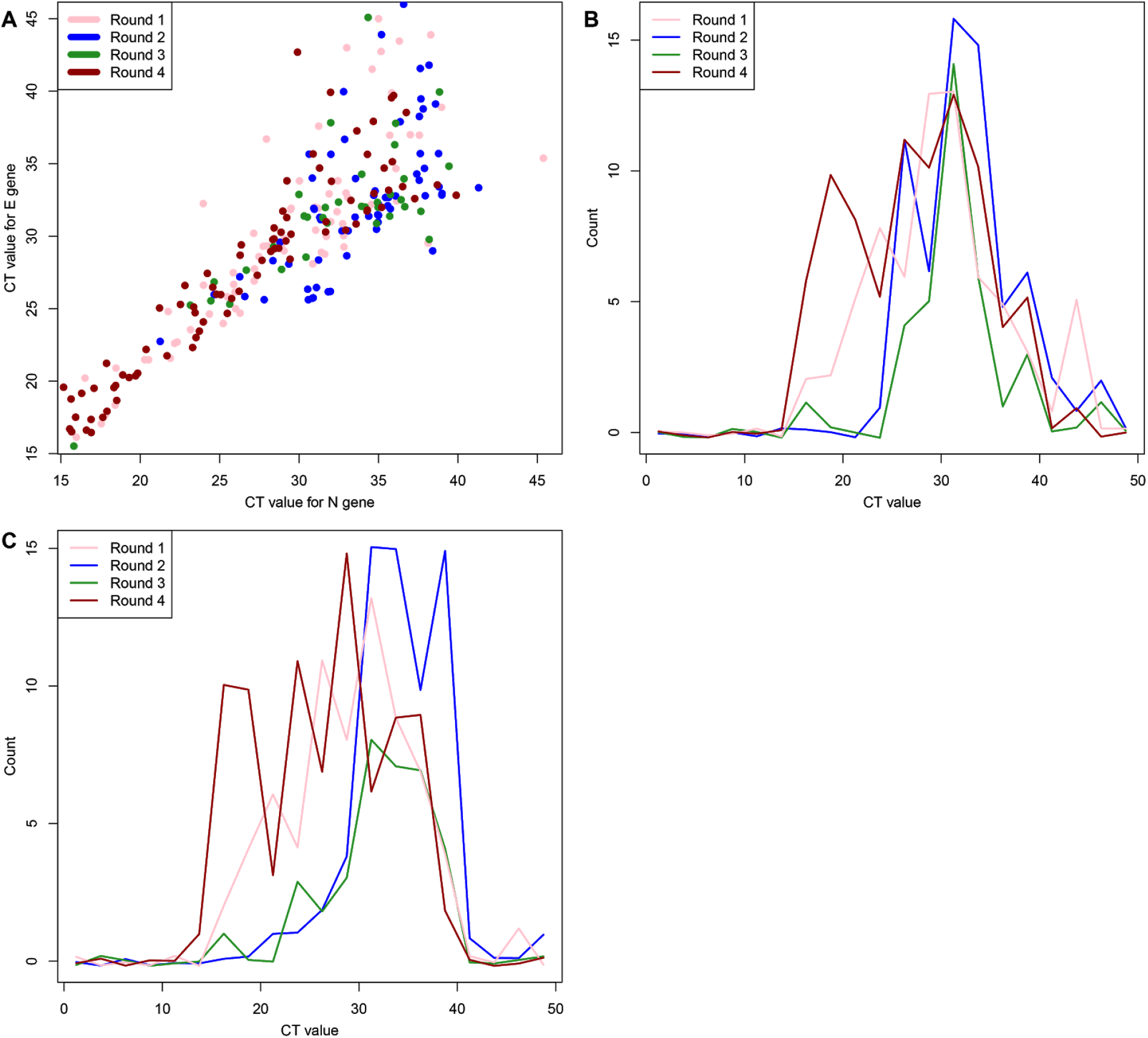
Comparison of cycle threshold (CT) values between rounds for samples in which both E and N genes were detected. **A** scatter plot of CT values for rounds 1 (pink), 2 (blue), 3 (green) and 4 (brown). **B** (E gene) and **C** (N gene) histograms of CT values for rounds 1, 2, 3 and 4.

**Table 7.** Comparison of cycle threshold (CT) values between rounds of the REACT-1 study.

| Round | CT values |  |  | Between round p-value |  |  |  |
| --- | --- | --- | --- | --- | --- | --- | --- |
|  | Median | Min | Max | Round 1 | Round 2 | Round 3 | Round 4 |
| 1 | 29.5 | 16.0 | 45.4 | - | 0.001 | 0.211 | 0.514 |
| 2 | 34.8 | 21.3 | 47.6 | 0.001 | - | 0.293 | < 0.001 |
| 3 | 33.9 | 15.8 | 39.4 | 0.211 | 0.293 | - | 0.021 |
| 4 | 27.4 | 14.5 | 39.9 | 0.514 | < 0.001 | 0.021 | - |

We tested for the presence of clustering at arbitrary spatial scale, independent of geographical units, by combining data in sequential rounds. We identified clusters in and around London, transient clusters in the Midlands, and an expanding area of clustering in the North West and, more recently, in Yorkshire and the Humber (Figure 5). The spatial pattern of most clustered swab-positive cases was robustly different from 10,000 random draws of the most clustered samples of negative cases (Figure 6).

**Figure 5.**
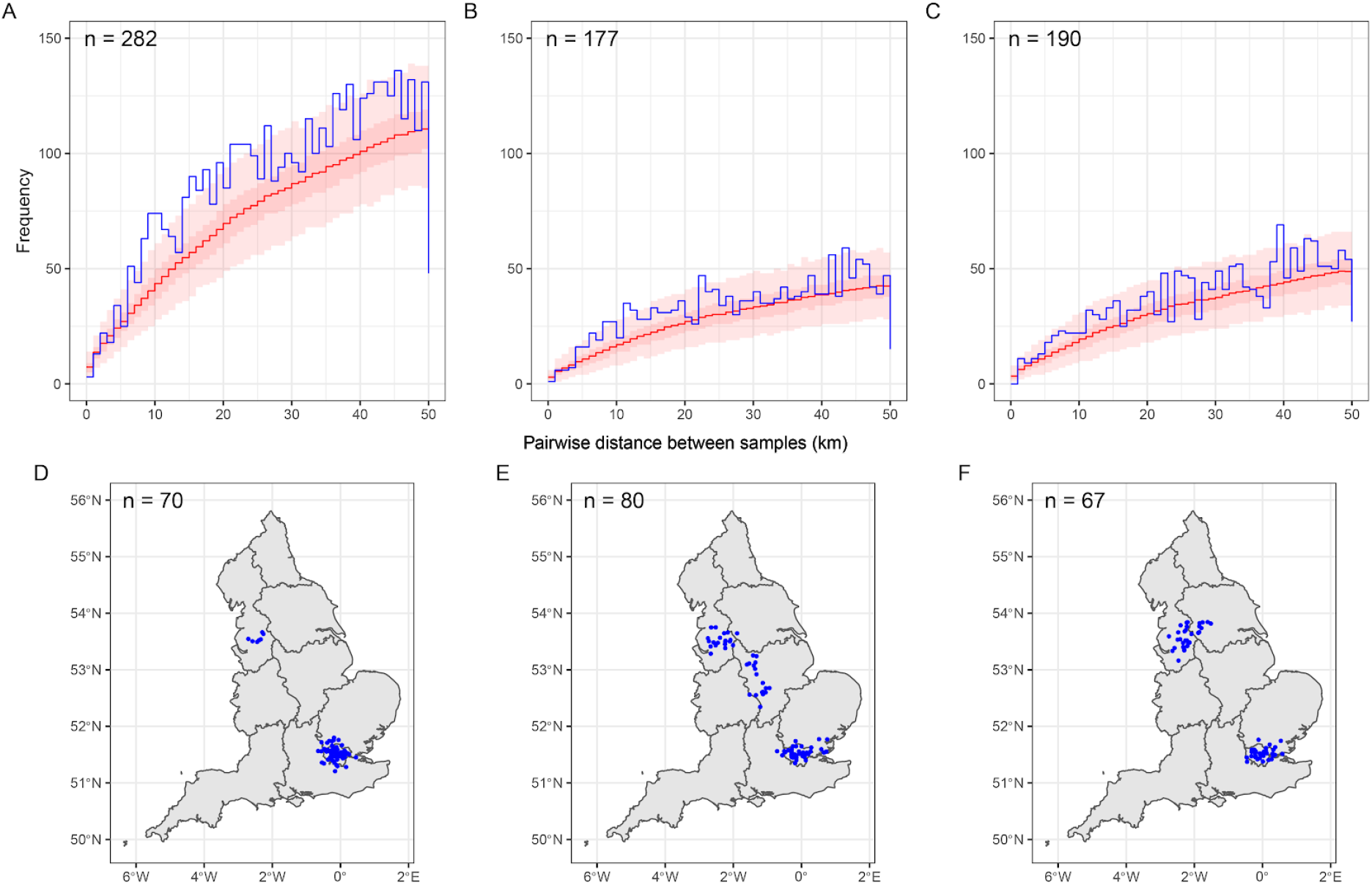
Spatial clustering during sequential rounds of REACT-1. **A** (rounds 1 and 2), **B** (rounds 2 and 3), **C** (rounds 3 and 4) the distribution of pairwise distances between swab-positive participants (blue line) compared with 5000 random redraws of pairwise distances between swab-negative participants (red line, median; dark red area, central 50%; light red area, central 95%). **D** (rounds 1 and 2) and **E** (rounds 2 and 3) **F** (rounds 3 and 4) jittered home locations of swab-positive participants who most frequently formed close pairs with other positive participants (see Figure 6 and Methods).

**Figure 6.**
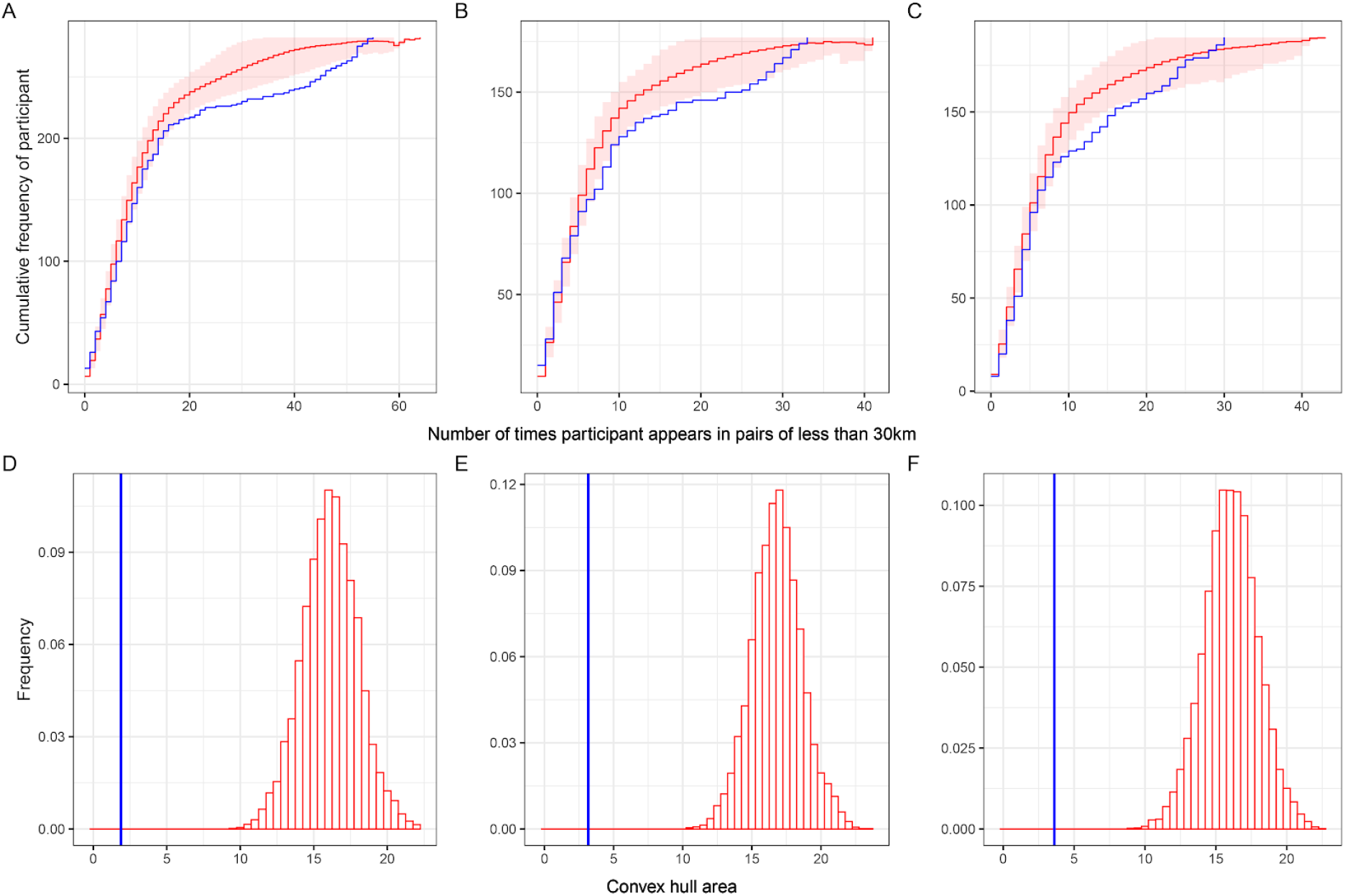
Identifying highly clustered positive samples. Cumulative distribution of the number of times participants with positive (blue) or negative (red) tests are in pairs of distance 30 km apart or less: (**A**) rounds 1 and 2, (**B**) rounds 2 and 3, (**C**) rounds 3 and 4. Red line averaged over 5000 random draws of 282 negative participants. Red area is the central 90% range. Distribution of areas of convex hulls (red bars) around home locations of most clustered negative participants in rounds 1 and 2 (**D**), rounds 2 and 3 (**E**), and rounds 3 and 4 (**F**) from 10 000 repeated random draws. Vertical blue line shows area of convex hull of the same number of most clustered positive participants.

## Discussion

We demonstrate that our national community surveillance programme based on the experience of over 600,000 randomly tested individuals across England was able to quantify a fall and then give early warning of a rise in prevalence between May and early September 2020. Our estimate of prevalence during round 4 in August-September now exceeds that which we reported for June and July 2020 but is still well below those seen during the peak of the epidemic in March and April 2020. Unlike testing programmes for symptomatic individuals being rolled out in many countries [12], our study has the advantage that it is not biased by the propensity to seek testing or by service capacity.

Our findings show important variations by age. In our most recent data, there is evidence of increasing prevalence in younger age groups, particularly those aged 18 to 24 years. However, prevalence appears also to be increasing among older adults, up to the age of 65, suggesting that the resurgence of SARS-CoV-2 transmission in England is already affecting at-risk populations [13].

At a regional level, initially the highest prevalence was recorded in London, which declined across rounds 1 to 3 before rising again in round 4. There has been a marked recent rise in prevalence in the north of England (North West, North East and Yorkshire and The Humber) such that these regions now have the highest prevalence nationally. However, a rise in prevalence is not restricted to these regions and can be seen across all areas. We also found evidence for clustering of cases at the sub-regional level, which developed over time between regions. Initially we reported clustering of cases in and around London, which may reflect patterns of commuting by train into a large city [14]. Over time, while clustering in and around London remained, we saw local clusters emerging in the North West, East Midlands and, more recently, in Yorkshire and The Humber. However, clustering detected in East Midlands was no longer apparent in the most recent data, which may partly reflect successful local lockdowns in that region.

We find differences over time in the odds of infection for health care and care home workers. During round 1 (May) such workers had over five-fold odds of infection compared with non-key workers, reflecting evidence of transmission in both health care and care home settings. However, the increased odds declined in subsequent rounds. It is apparent that there has been a shift in transmission from hospital and care home settings earlier in the epidemic to community transmission.

Across the four rounds of our study, we found that people of Asian ethnicity (mainly South Asian) had consistently higher odds of infection than white people, around 2-fold higher in the most recent data. This higher rate of infection is also reflected in higher SARS-CoV-2 seroprevalence among people of Asian ethnicity in England [15]. These data suggest that higher rates of hospitalisation and mortality from COVID-19 reported amongst people of Asian ethnicity in England may be explained by higher rates of infection, rather than by a predisposition to progress to severe disease once infected. Our results also suggested higher odds of infection compared to whites among people of Black, mixed and other ethnicities.

Our study has limitations. First, although we aimed to be representative of the population of England by inviting a random sample of people on the NHS patient register, differential response by e.g. age, sex, ethnicity may have distorted our findings. For overall estimates of prevalence we reweighted our sample to be representative of England taking into account sample design and non-response. Prevalence estimates for subgroups were not reweighted because of low numbers of swab-positive participants.

Second, we relied on self-swabbing to obtain estimates of swab positivity. A throat and nose swab is estimated to have 70% to 80% sensitivity [16], so we are likely to have underestimated true prevalence, although, this would not be expected to have affected trend analysis or estimation of the R value.

Third, it is possible that at least part of the trends we observe in swab positivity during our study may have been the result of changing availability symptom-driven test capacity. We mitigated this potential bias by monitoring the proportion of swab-positive participants who were asymptomatic (overall 72%, ranging by round from 65% to 81%) and by repeating R estimates for the subset of asymptomatic individuals in each round.

Our findings have implications for control of the COVID-19 pandemic. Until effective vaccines are available and widely disseminated, control of the SARS-CoV-2 virus must rely on established public health measures [17] including social distancing, frequent hand-washing, and face covers. Our early data, as we exited lockdown, demonstrate the high level of effectiveness of stringent social distancing in reducing transmission of the virus, with prevalence rates decreasing by 75% over a 3 month period to early August. However, since then prevalence has increased, perhaps reflecting holiday travel, return to work, or a more general increase in the number and transmission potential of social interactions. While in England there has yet to be notable increases in hospitalisations or deaths associated with the resurgence in infection, this is not the case in other European countries, such as France and Spain, where hospitalisations are increasing substantially [1].

By detecting a resurgence of SARS-CoV-2 transmission in the community in England at an early stage, there is an opportunity to intervene while prevalence is still at a relatively low level. The objective for ongoing public health policy should be to limit the spread of the virus such that R drops below one, while maintaining as much social interaction and economic activity as possible. Studies like REACT-1 are needed not only as an early warning system, but also to help rapidly evaluate the success of such policies in keeping the virus controlled at low prevalence.

## Data Availability

The datasets generated or analysed, or both, during this study are not publicly available because of governance restrictions.

## Acknowledgements

SR, CAD acknowledge support: MRC Centre for Global Infectious Disease Analysis, National Institute for Health Research (NIHR) Health Protection Research Unit (HPRU), Wellcome Trust (200861/Z/16/Z, 200187/Z/15/Z), and Centres for Disease Control and Prevention (US, U01CK0005-01-02). GC is supported by an NIHR Professorship. PE is Director of the MRC Centre for Environment and Health (MR/L01341X/1, MR/S019669/1). PE acknowledges support from the NIHR Imperial Biomedical Research Centre and the NIHR HPRUs in Environmental Exposures and Health and Chemical and Radiation Threats and Hazards, the British Heart Foundation Centre for Research Excellence at Imperial College London (RE/18/4/34215) and the UK Dementia Research Institute at Imperial (MC_PC_17114). We thank The Huo Family Foundation for their support of our work on COVID-19.

We thank key collaborators on this work -- Ipsos MORI: Kelly Beaver, Sam Clemens, Gary Welch, Andrew Cleary and Kelly Ward; Institute of Global Health Innovation at Imperial College: Gianluca Fontana, Dr Hutan Ashrafian, Sutha Satkunarajah and Lenny Naar; MRC Centre for Environment and Health, Imperial College London: Daniela Fecht; North West London Pathology and Public Health England for help in calibration of the laboratory analyses; NHS Digital for access to the NHS register; and the Department of Health and Social Care for logistic support. SR acknowledges helpful discussion with members of the UK Government Office for Science (GO-Science) Scientific Pandemic Influenza - Modelling (SPI-M) committee.

